# The safety and efficacy of using age-adjusted D-dimers in hospitalised patients in a diverse urban centre: a real-world data study

**DOI:** 10.1101/2024.06.02.24308329

**Authors:** S. Gallier, F. Evison, J. Hodson, R. Khosla, T. Ranasinghe, L. Rickard, C. Atkin, V. Reddy-Kolanu, K. Nirantharakumar, W. Lester, B. Holloway, E. Sapey

## Abstract

**Objective:** Despite recommendations, age adjusted thresholds (AAT) for D Dimers are not routinely used as part of venous thromboembolism (VTE) screening in many healthcare settings due to concerns about missing cases, especially in older and co-morbid adults. The National Institute for Health and Care Excellence in the UK has highlighted that evidence to support AAT is not plentiful. This study assessed the real-world use of AAT D-dimers for VTE in a large cohort of acutely hospitalised patients.

**Methods:** This retrospective data study included all adult patients attending a large hospital with a suspected VTE between January 2017 to December 2021. The predictive accuracy of D-dimer was assessed against gold standard imaging. Outcomes of false negative (with AAT) and false positives (with standard thresholds) cases were assessed.

**Results:** 27,526 suspected VTE attendances were included, with a 4.3% confirmed VTE diagnosis rate. The ST D-dimer exhibited high sensitivity (91.1%) but modest specificity (65.2%). The AAT demonstrated slightly lower sensitivity (87.0%) but higher specificity (71.7%, p<0.001). The performance of ST thresholds declined with age, with false positive rates increasing from 17.4% to 80.0% in people aged < 50 years and > 90 years respectively. The AAT accurately identified 1,700 true negatives misclassified as false positives by the ST. 14 patients in this group were admitted with a bleed within 30 days. AAT misdiagnosed 24 cases as false negatives, with most being small sub-segmental pulmonary emboli or non-occlusive DVTs. Using AAT thresholds could have avoided 64 scans per 1,000 attendances, saving approximately £235,310 of imaging costs in this cohort.

**Conclusion:** The age-adjusted D-dimer threshold enhances diagnostic precision and could decrease unnecessary imaging and anticoagulation, reducing investigations with time and cost savings with no significant safety signal.

## Background

Venous thromboembolic events (VTE), presenting as deep vein thrombosis (DVT) and pulmonary embolism (PE), impact millions of people worldwide each year ^(1–3)^, ranking as the third leading cause of acute cardiovascular syndrome ^(4, 5)^. In the UK, the incidence rate is 1-2 per 1,000 people ^(6, 7)^ and the crude mortality rate associated with VTE within 90 days following discharge from hospital is 99.4 per 100,000 people ^(8)^. Due to an ageing and multi-morbid population, VTE events are increasingly common. Over 69,064 hospital episodes of PE were reported in the UK between 2021-2022 which resulted in 36,757 admissions ^(9)^.

With significant mortality and morbidity associated with this disease, it is vital to diagnose VTE as early as possible. However, VTE diagnosis is complicated by the non-specific nature of its presenting symptoms and the high frequency by which these symptoms occur ^(10)^. There are clinical scores which aid the diagnosis of VTE by stratifying people into low or high probability groups. In those with a low probability, a plasma D-dimer test can be used to rule out disease. In those with a raised D-dimer or a high probability of VTE, definitive medical imaging is required. In the UK, the National Institute for Health and Care Excellence (NICE) support the use of the Wells score with a D-dimer test ^(11, 12)^ and computed tomography pulmonary angiography (CTPA) or ventilation-perfusion scan (VQ) for a suspected PE or ultrasound scans (USS) for a suspected DVT ^(13)^. This approach has a very low rate of diagnostic failure ^(14–16)^. However, a large increase in CTPAs and ultrasounds has been seen for suspected PE/DVT, placing a significant burden on diagnostic services ^(17, 18)^.

Initial risk prediction models used a standardised threshold for a D-dimer test (ST), but it was increasingly recognised that D-dimers can become elevated with age and in several other systemic conditions. Several studies have indicated an increased specificity in diagnosing DVT or PE when utilising age-adjusted D-dimer threshold (AAT)^(19–22)^including systematic reviews ^(23, 24)^. The 2020 NICE guideline [NG158] for the diagnosis of VTE suggested that clinicians consider utilising AAT for people aged over 50 years of age ^(25)^. Despite this, the adoption of AAT D-dimers remains limited across healthcare organisations. Reasons for this include concerns regarding the accuracy of AAT D-dimers in small studies with not all studies showing benefit, clinical uncertainty about the use of AAT D-dimers across diverse populations, and a lack of information on the potential clinical consequence of false negatives using an AAT approach^(26–28)^.

This retrospective data study aimed to assess and compare the predictive accuracy of ST and AAT D-dimer in patients presenting acutely to hospital with suspected VTE. Separate analyses were also performed for the outcomes of PE and DVT individually and across differing age groups and comorbidities. The secondary aim was to assess the characteristics and presentations of patients who might be misdiagnosed comparing an AAT to a ST for D-dimer. This included an individual note review of all false positives and negatives using an AAT to determine the clinical significance of missing a VTE or providing treatment for a VTE when no such VTE was present. The final aim was to calculate the potential impact on service utilisation including imaging requests (namely CTPA/VQ and Ultrasound tests) were an age-adjusted approach applied.

## Methods

The study was supported by PIONEER, a Health Data Research Hub in Acute Care. Ethical approvals for the study were provided by the East Midlands – Derby REC (reference: 20/EM/0158). Two in-depth reviews of identifiable patient notes were conducted as part of a service evaluation, both approved by University Hospitals Birmingham NHS Foundation Trust (UHB) Service Evaluation and Clinical Audit Team. In the first, there was a false negative using an AAT D-dimer but a true positive using an ST D-dimer (reference CARMS-21061). The second assessed anyone readmitted with a haemorrhage following a D-dimer in the false positive ST cohort (reference CARMS-21227).

### Setting

The study was based on retrospective data collected from the electronic heath record (EHR) system at Queen Elizabeth Hospital Birmingham (QEHB), part of UHB, one of the largest NHS Trusts in England. Patients either presented to the emergency department (ED) or were referred by a general practitioner or other clinical service (e.g. the UK’s 111 system) to the acute medical assessment unit directly. If VTE was suspected on triage, risk stratification was performed by the internal medicine or emergency medicine team.

### Study cohort

The primary inclusion criteria for the study were patients attending QEHB between 1^st^ Jan 2017 and 31^st^ Dec 2021, with a suspected diagnosis of VTE and where a D-dimer test was performed for that suspected VTE. The following exclusion criteria were then applied, with further details provided in **Supplementary Figure 1**:

- Patients who were taking anticoagulants at the time of the D-dimer test, as this impacts D-dimer interpretation ^(29)^. These were defined as patients who either reported having an active prescription for an anticoagulant at the time of attendance or who were administered a treatment dose of an anticoagulant less than 48 hours prior to the D-dimer test being performed.
- Re-attendances by the same patient within 90 days of the index attendance, as these likely represented the same underlying instance of suspected VTE.
- Patients aged <18 years at the time of attendance.

### Data collection

Data were retrospectively extracted from the EHR system at QEHB. The D-dimer level closest to the time of attendance was included, within a maximum interval of ±10 days. D-dimer tests reported levels in D-dimer units (DDU), with a lower limit of detection of 150μg/L. Any values below this threshold were assigned a value of 150μg/L for analysis and are reported as “<150μg/L”.

Dichotomisation of D-dimer levels used two different approaches: a standard threshold (ST) value of 250μg/L ^(24, 30)^, and an age-adjusted threshold (AAT), which used a value of 250μg/L for those aged <50 years, or age (in years) x 5μg/L for those older than 50 years ^(31–33)^.

Baseline characteristics were extracted, including age, sex, ethnicity, and COVID-19 status. Deprivation was quantified using the index of multiple deprivation (IMD), which was categorised based on national quintiles for analysis ^(34)^. The BMI measurement recorded closest to the index attendance, within ±6 months, was also extracted. The primary presenting complaint was identified based on the details recorded on arrival at the ED; this was not routinely available for patients who directly attended medical or surgical wards, including those referred by their GP, and those on the community DVT pathway. The presence of underlying comorbidities at attendance was identified based on the ICD10 codes recorded at discharge. The first Wells’ scores for either PE (Wells-PE) or DVT (Wells-DVT) performed either during the index attendance or a follow-up attendance to the specialist PE/DVT clinic were also extracted, where available, as was the first NEWS2 score recorded during the index attendance.

The primary outcome was the diagnosis of VTE, which was a composite of DVT and PE. The outcome was defined as the presence of an associated ICD10 code either during index attendance, or within ten or five days of discharge for DVT and PE, respectively. ICD10 codes used were: *I80.1, I80.2, I80.3, I80.9, O22.3,* and *O87.1* for DVT, and *I26.X* for PE. Additional outcomes included the total hospital length of stay, mortality during the index attendance, and at three-, six- and twelve-months post-discharge.

Two service evaluations were conducted on those people with a confirmed VTE diagnosis who met the ST but not AAT D-dimer threshold (a D-dimer level of >250μg/L but less than the AAT) – AAT false negatives. For each of these patients, medical notes and imaging review was undertaken by a consultant radiologist and consultant medical physician in an MDT. For each VTE “missed” by AAT, clinicians rated the VTE as low (sub-segmental PE or nonocclusive DVT), or high risk (multiple sub-segmental or segmental PE or occlusive DVT). Two medical consultants also conducted a service evaluation that reviewed medical notes and discharge summaries of patients who had a raised ST D-dimer which was below the AAT threshold (ST false positives) especially assessing risks associated with anti-coagulation where this was given while awaiting definitive imaging (up to 7 days). Here, any side effects were ranked as being unlikely to be caused by anticoagulation, potentially caused by anticoagulation or highly likely to be caused by anticoagulation, according to the MDT.

### Statistical methods

The predictive accuracy of D-dimer with respect to VTE was initially quantified using the area under the receiver operating characteristic curve (AUROC). The classification accuracies of the ST and AAT were then quantified using a range of measures of test performance, which were compared between the two thresholds using Fisher’s exact test. This analysis was also repeated within subgroups of age, with trends in sensitivity and specificity visualised using binary logistic regression models with age as a continuous covariate. All analyses were performed using IBM SPSS 24 (IBM Corp. Armonk, NY), with p<0.05 deemed to be indicative of statistical significance throughout. Cases with missing data were excluded from the analysis of the affected variable, unless stated otherwise. Continuous variables were not found to follow normal distributions, and so are summarised using medians and interquartile ranges (IQRs) throughout.

Furthermore, the study assessed the number of imaging scans, specifically CTPA and USS, that could have been circumvented utilising the AAT along with the subsequent potential cost savings. VQ were also identified, for those patients with contraindications to CTPA, and were combined with CTPA scans for analysis including cost modelling. The direct access costs of imaging patients were taken from the latest NHS Reference Costs 2021/22, which were last updated May 2023(**^35^**). The direct access cost for a CTPA is £122.87 and £85.21 for a USS. The number of return visits for care completion in ST false positives were recorded.

## Results

### Cohort characteristics

A total of N=27,526 attendances of patients with suspected VTE met the inclusion criteria of the study (see **Supplementary Figure 1** for study flowchart). Patients had a median age of 53 years (IQR: 37-69), with 57.7% female and 70.1% of White ethnicity; the most common presenting complaint was chest pain (36.3%, **Table 1**). Wells-PE scores were only recorded in the structured EHR system for 14.4% of cases, with the Wells-DVT available for <0.1%; as such, Wells scores were not included in subsequent analysis. PE was diagnosed in N=693 (2.5%) cases and DVT in N=528 (1.9%), of whom N=41 had diagnoses of both PE and DVT. As such, the composite outcome of VTE diagnosis occurred in N=1,180 (4.3%) cases. The in-hospital mortality rate was 2.0%, rising to 8.1% within 12 months post-discharge.

**Table 1.**
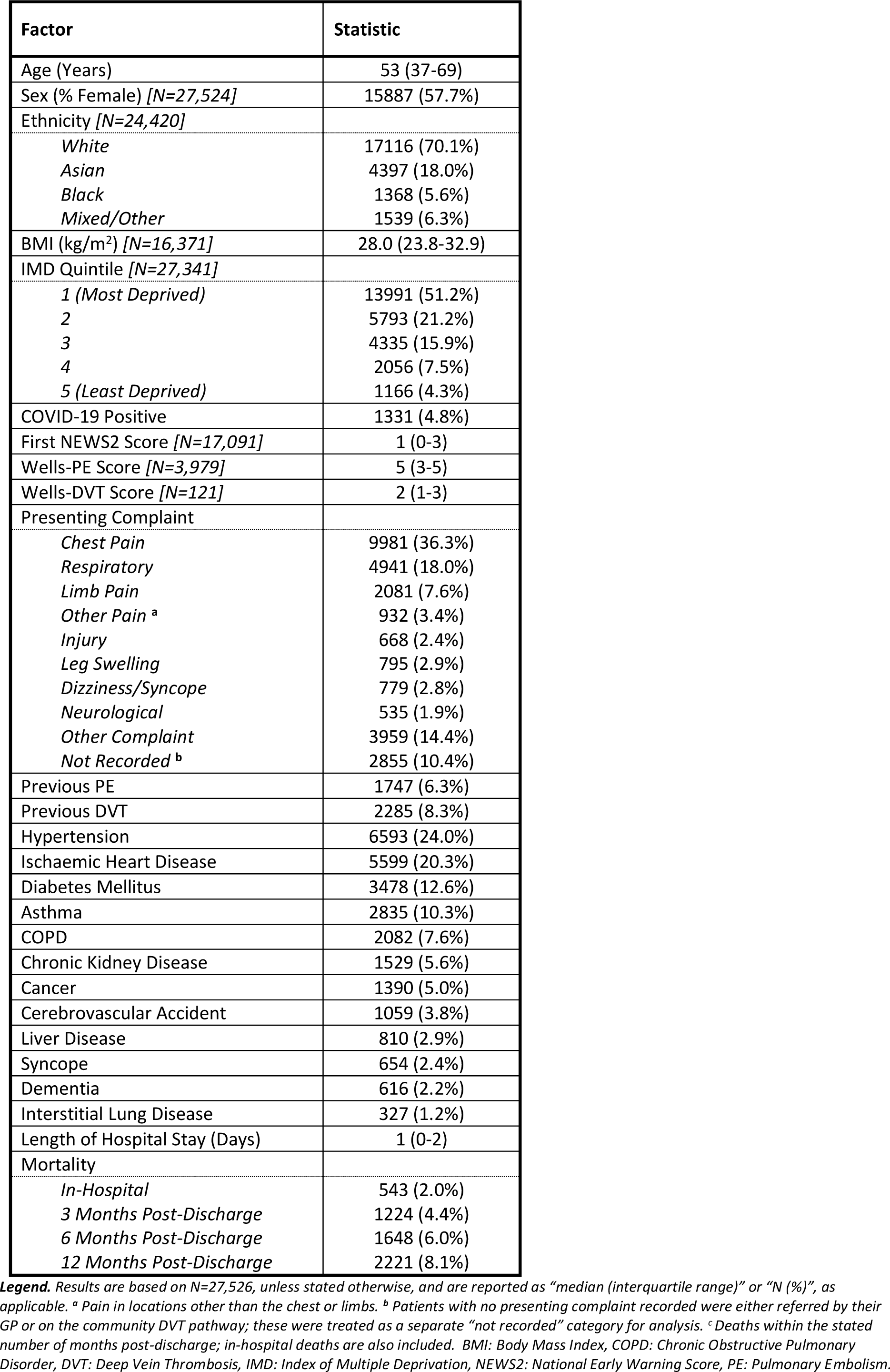
Cohort characteristics.

### Predictive accuracy of D-dimer levels

The median D-dimer for the cohort was 178μg/L (IQR: <150-383), with 43.4% (N=11,937) having D-dimer values below the lower limit of detection of the assay (<150μg/L). D-dimer was found to be a strong predictor of VTE, with an AUROC of 0.863 (95% CI: 0.835-0.874). When considering the components of the composite outcome separately, performance of D-dimer was superior for PE (AUROC: 0.898, 95% CI: 0.888-0.909) compared to DVT (0.804, 0.785-0.823).

### Classification accuracy of D-dimer thresholds

A total of 37.2% (N=10,244) of cases had D-dimer levels above the ST (≥250μg/L) and, hence, were classified as being at high risk of VTE based on this threshold. Of these, N=1,748 had D-dimer levels that were below the AAT; hence, would have been reclassified as low risk had the AAT been used instead of the ST (**Figure 1**). These comprised N=1,700 cases who were not diagnosed with VTE; hence, represented additional true negatives for the AAT. However, the remaining N=48 patients were diagnosed with VTE; hence, would have been incorrectly deemed low risk by the AAT, and represented additional false negatives.

**Figure 1.**
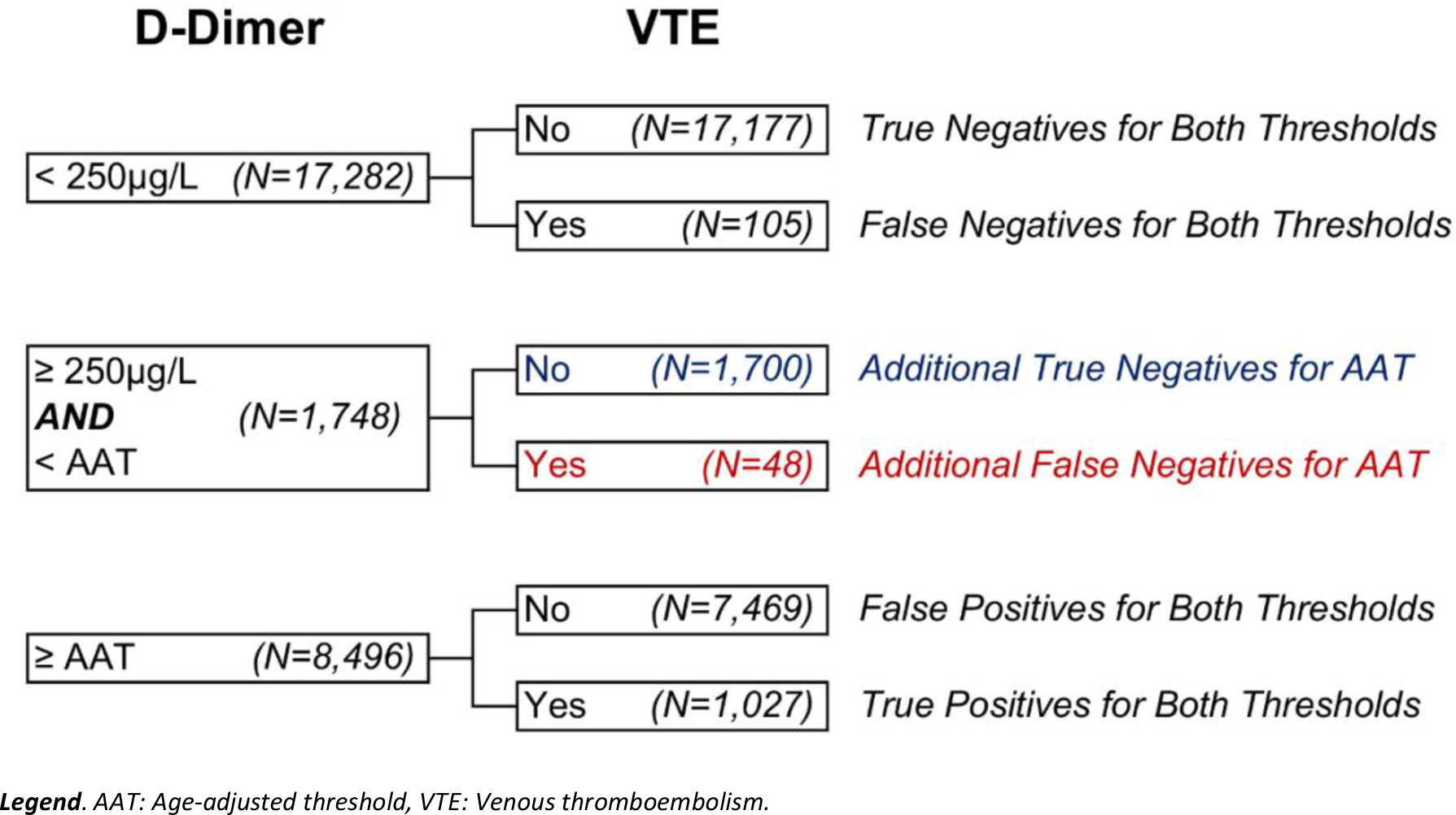
Flowchart of VTE diagnoses by D-dimer threshold.

To assess the impact of these discrepancies between the two thresholds, the classification accuracies of both thresholds, with respect to VTE, were then compared (**Table 2**). This found the ST to have sensitivity of 91.1%, with a negative predictive value (NPV) of 99.4%. However, specificity was modest at 65.2%, with only 10.5% of cases with D-dimer levels above the ST being diagnosed with VTE. The AAT had a significantly lower sensitivity (87.0% vs. 91.1%, p=0.002) and NPV (99.2% vs. 99.4%, p=0.028) than the ST, as a result of the additional N=48 false negatives. However, the AAT also had a significantly higher specificity (71.7% vs. 65.2%, p<0.001).

**Table 2.** Classification accuracy of D-dimer thresholds.

| D-dimer Threshold | Outcome |  | Sensitivity | Specificity | Negative Predictive Value | Positive Predictive Value | Accuracy | Odds Ratio (95% CI) |
| --- | --- | --- | --- | --- | --- | --- | --- | --- |
|  | No | Yes |  |  |  |  |  |  |
| **Outcome = VTE** |  |  | ***p=0.002*** | ***p<0.001*** | ***p=0.028*** | ***p=0.001*** | ***p<0.001*** | - |
| Standard Threshold |  |  |  |  |  |  |  |  |
| <250µg/L | 17177 | 105 | 91.1% | 65.2% | 99.4% | 10.5% | 66.3% | 19.2 |
| ≥250µg/L | 9169 | 1075 | (1075/1180) | (17177/26346) | (17177/17282) | (1075/10244) | (18252/27526) | (15.6-23.5) |
| Age-Adjusted Threshold |  |  |  |  |  |  |  |  |
| <Threshold | 18877 | 153 | 87.0% | 71.7% | 99.2% | 12.1% (1027/8496) | 72.3% | 17.0 |
| ≥Threshold | 7469 | 1027 | (1027/1180) | (18877/26346) | (18877/19030) |  | (19904/27526) | (14.3-20.1) |
| **Outcome = DVT** |  |  | ***p=0.045*** | ***p<0.001*** | ***p=0.244*** | ***p=0.059*** | ***p<0.001*** | - |
| Standard Threshold |  |  |  |  |  |  |  |  |
| <250µg/L | 17200 | 82 | 84.5% | 63.7% | 99.5% | 4.4% | 64.1% | 9.5 |
| ≥250µg/L | 9798 | 446 | (446/528) | (17200/26998) | (17200/17282) | (446/10244) | (17646/27526) | (7.5-12.1) |
| Age-Adjusted Threshold |  |  |  |  |  |  |  |  |
| <Threshold | 18922 | 108 | 79.5% | 70.1% | 99.4% | 4.9% | 70.3% | 9.1 |
| ≥Threshold | 8076 | 420 | (420/528) | (18922/26998) | (18922/19030) | (420/8496) | (19342/27526) | (7.4-11.3) |
| **Outcome = PE** |  |  | ***p=0.012*** | ***p<0.001*** | ***p=0.036*** | ***p=0.004*** | ***p<0.001*** | - |
| Standard Threshold |  |  |  |  |  |  |  |  |
| <250µg/L | 17256 | 26 | 96.2% | 64.3% | 99.8% | 6.5% | 65.1% | 46.2 |
| ≥250µg/L | 9577 | 667 | (667/693) | (17256/26833) | (17256/17282) | (667/10244) | (17923/27526) | (31.2-68.4) |
| Age-Adjusted Threshold |  |  |  |  |  |  |  |  |
| <Threshold | 18982 | 48 | 93.1% | 70.7% | 99.7% | 7.6% | 71.3% | 32.5 |
| ≥Threshold | 7851 | 645 | (645/693) | (18982/26833) | (18982/19030) | (645/8496) | (19627/27526) | (24.2-43.6) |

**Legend.** The age-adjusted threshold used a value of 250µg/L for those aged <50 years, or age (in years)\*5µg/L for those older than 50 years. *p*-values are from Fisher's exact tests, comparing the percentages between the standard and age-adjusted thresholds; bold *p*-values are significant at *p*<0.05. CI: confidence interval, DVT: Deep Vein Thrombosis, PE: Pulmonary Embolism, VTE: Venous thromboembolism.

### Impact of patient age on classification accuracy

D-dimer levels were found to increase significantly with age (p<0.001), with this effect being most pronounced in those that were not diagnosed with VTE. Specifically, for patients without a VTE diagnosis, the median D-dimer increased from <150μg/L (IQR: <150-195) in those aged <30 years to 480μg/L (IQR: 282-852) in those aged 90+ years, with a corresponding increase in the proportion with D-dimer levels above the standard threshold from 17.4% to 80.0% (**Figure 2**).

**Figure 2.**
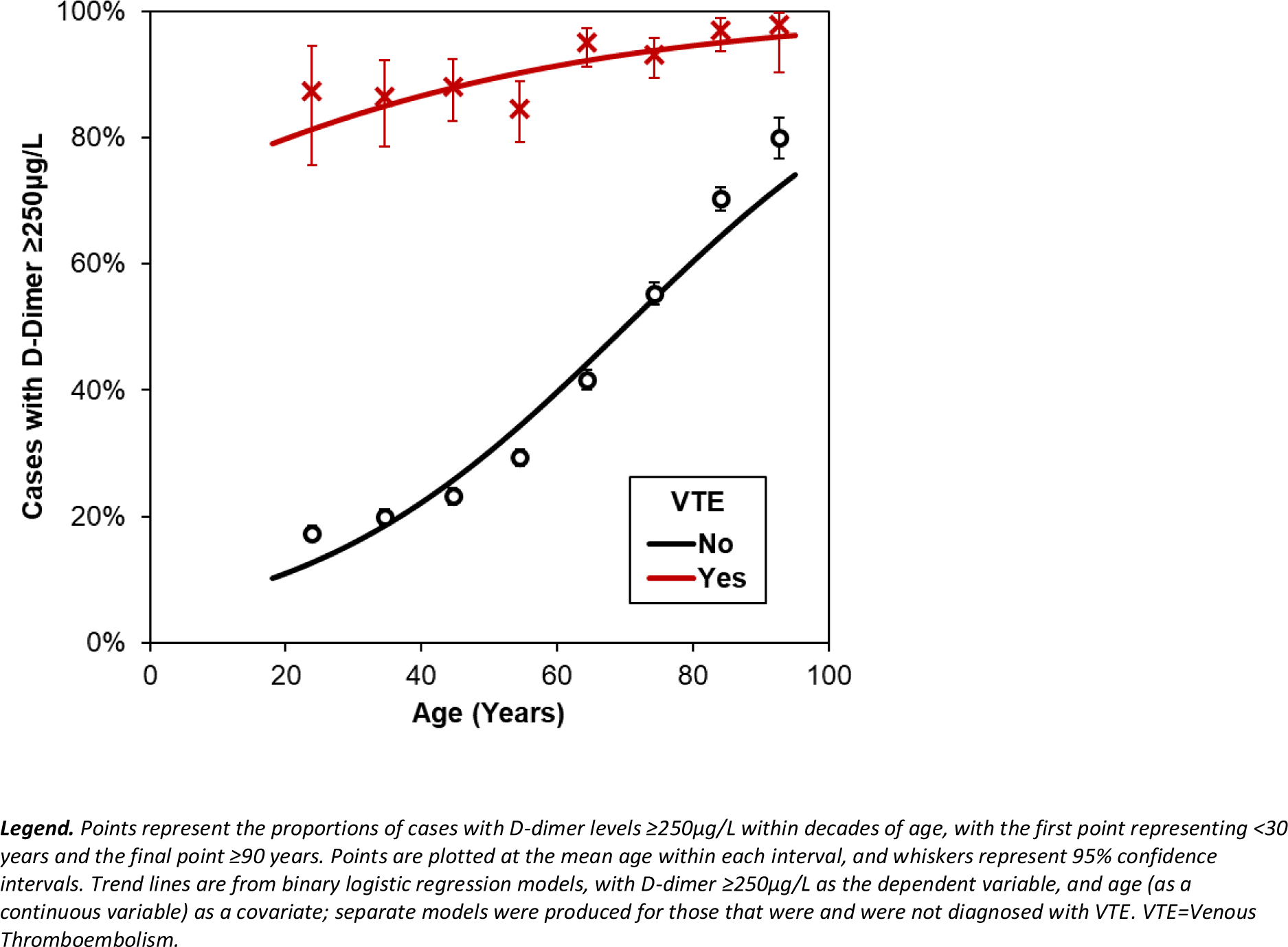
Association between age and the likelihood of D-dimer ≥250μg/L.

In light of this, the effect of age on the classification accuracy of the two thresholds was then assessed. This found a similar trend for both thresholds, with increasing age associated with progressive increases in sensitivity with a corresponding reduction in specificity (**Figure 3, Supplementary Figure 1**). However, this effect was more pronounced for the ST, leading to the difference in classification accuracy between the two thresholds increasing with age. For example, despite both thresholds having identical performance in patients ≤50 years (as the ST and AAT were both 250μg/L in this range), for patients aged 80-89 the AAT had significantly lower sensitivity (86.8% vs. 97.0%, p=0.001) but significantly higher specificity (54.4% vs. 29.7%, p<0.001) than the ST.

**Figure 3.**
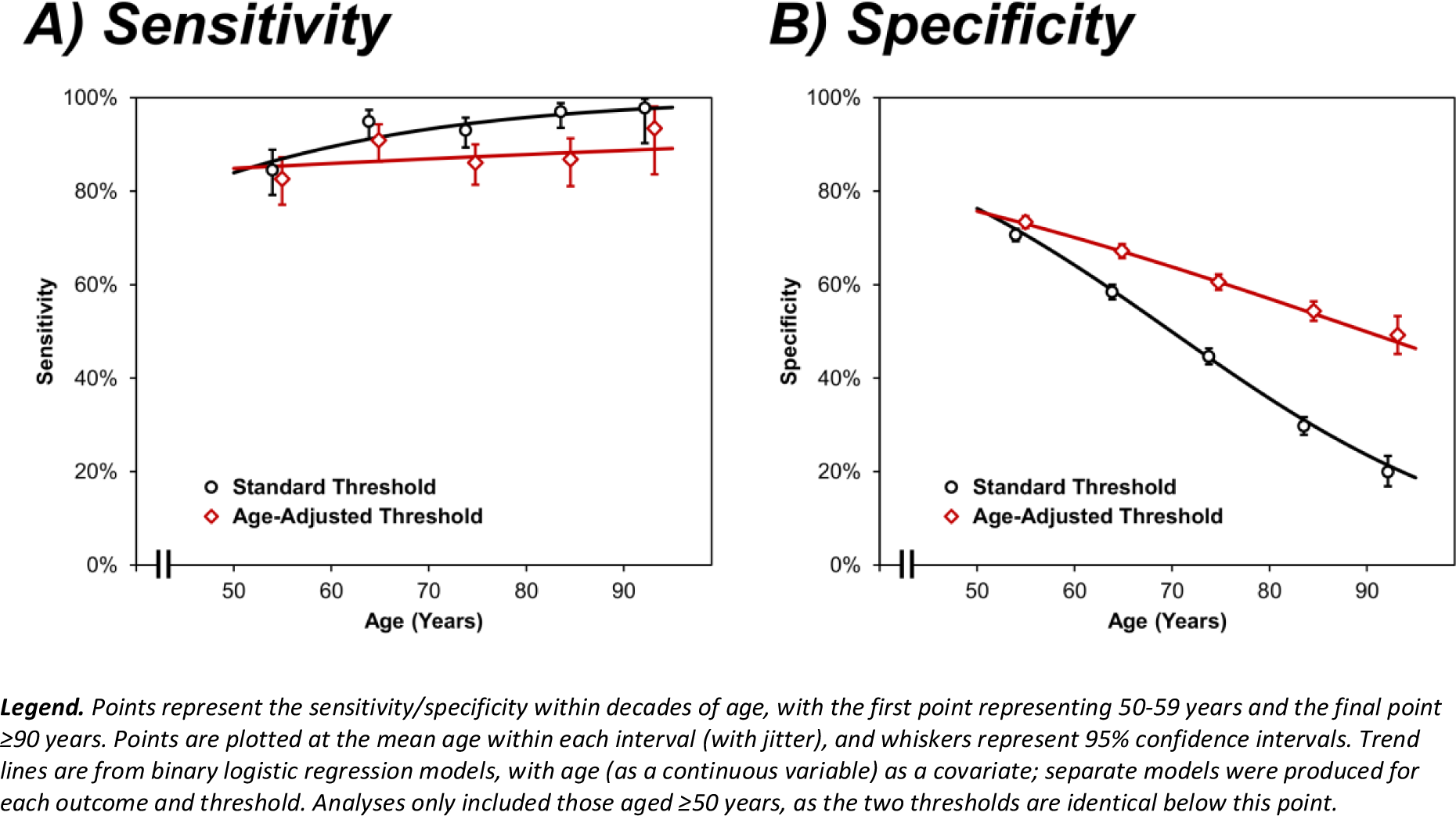
Sensitivity and specificity of D-dimer thresholds by age.

The analysis of the classification accuracy of the ST and AAT was then repeated for the subgroups of patients with comorbidities (**Supplementary Table 3**). This returned similar results to the analysis of the cohort as a whole. Specifically, the AAT had significantly greater accuracy and specificity than the ST for all of the subgroups considered. There was also a tendency for the AAT to have lower sensitivity and higher PPV, although these comparisons generally did not reach statistical significance, largely due to the small within-group sample sizes leading to insufficient statistical power.

### Review of the additional false negatives associated with the AAT D-dimer

The AAT yielded an extra N=48 false negatives (0.17% of the overall cohort) compared to the ST, namely cases with VTE diagnoses who had been correctly identified as high risk by the ST, but deemed low risk by the AAT; these cases were reviewed as part of a service evaluation. Of these, N=24 had been diagnosed with VTE based on initial imaging which was suggestive of VTE, but inconclusive; follow-up imaging in these patients found no evidence of VTE. As such, it is likely that only the remaining N=24 represented genuine cases of VTE, which is equivalent of an additional 0.9 false negatives per 1,000 attendances for the AAT. Of these patients, N=17 were diagnosed with PEs and N=7 with DVTs; where sufficient imaging was available (N=15), all PEs were small and sub-segmental or inconclusive/sub-optimal imaging, with all DVTs being partial venous occlusions behind the knee. Of note, only N=3 of these cases commenced anticoagulation during the admission, with this potentially being due to a second indication (atrial fibrillation) in N=2. There were N=2 in-hospital deaths; however, neither of these appeared to be related to VTE, instead being attributed to bowel perforation related to malignancy and congestive cardiac failure related to ischaemic heart disease, respectively.

### Review of the additional true negatives associated with the AAT D-dimer

The AAT yielded an extra N=1,700 true negatives (6.2% of the overall cohort) compared to the ST, namely cases without VTE who had been identified as high risk by the ST, but deemed low risk by the AAT. As such, these patients had the potential to be overtreated, with the associated risk of haemorrhage. Anticoagulation was prescribed in 65.3% (N=1,079) of this subgroup, with 79.5% (N=1,351) being admitted to a hospital ward, who had a median subsequent hospital stay of 51 (IQR: 8-166) hours. After leaving hospital, 0.8% (N=14) of patients had a subsequent attendance with a haemorrhage within 30 days; rates were similar in those that did and did not receive anticoagulation at the index admission (0.8% vs. 0.8%, p=1.000). A clinical service evaluation of these N=14 readmitted patients found that N=9 had anticoagulation prescribed, but this was only administered during the inpatient stay in N=4. Haemorrhage types included gastrointestinal bleeding, hemarthrosis, and vaginal bleeding; clinical review concluded that these haemorrhagic events were not related to anticoagulation therapy, due to the timing of the event compared to the administration of the medication.

### Change in imaging burden associated with the AAT D-dimer

If the decision to refer patients for imaging were made using a D-dimer threshold, then changing from the ST to the AAT could have reduced the number of cases referred for imaging by N=1,748 (i.e. the cases between the two thresholds), equivalent to 64 scans per 1,000 attendances. However, in practice, the D-dimer is not the only factor considered in clinical decision-making. Consequently, only N=658 (of N=1,748; 37.6%) of cases with D-dimer levels between the ST and AAT underwent imaging, comprising a total of N=393 CTPA or VQ scans and N=285 USS scans (N=20 had both scans), equivalent to 14 CTPA/VQ and 10 USS scans per 1,000 attendances. Based on these numbers and the NHS Reference Costs for the fiscal year 2021/22 (CTPA: £122.87, USS: £85.21), changing from the ST to the AAT would equate to a saving of £1,754 on CTPA/VQ and £882 on USS per 1,000 attendances.

## Discussion

Venous thromboembolism is a common cause of acute presentation to the hospital. The lack of sensitivity and specificity of signs and symptoms can make diagnosis challenging. Failure to diagnose VTE can result in dire outcomes, including sudden fatality, lasting cardiopulmonary complications, and a diminished quality of life ^(36)^. In patients with suspected VTE, distinguishing those without the condition is crucial to circumvent unnecessary anticoagulant treatment and its related haemorrhagic complications ^(37)^. Concurrently, excessive testing for PE can incur high costs and increase length of stay, adding to hospital crowding and amplifying service delivery pressures in already strained imaging departments, as well as subjecting patients to risks from radiation and IV contrast. There is evidence to support the use of AAT for D-dimers, and the theoretical benefits of this approach have been discussed, but the adoption of AAT remains patchy across health services.

This study, conducted on the largest, most diverse cohort to date, explored the efficacy of the age-adjusted threshold (AAT) compared to the standard threshold (ST) in D-dimer testing for diagnosing VTE. The findings offer important insights with potential implications for clinical practice, particularly in managing healthcare resources and improving diagnostic accuracy. The AAT showed a lower sensitivity (87.0%) than the ST (91.1%), indicating a slight increase in false negatives. The failure rate of 0.8% for AAT correlates to reported failure rates for USS that range between 0.57% and 2.0% (Cis ranging from 0.2% to 5.1%)^(38, 39)^. Compared to the reported venogram failure rate between 1.3% and 43.7%, it is reassuringly favourable ^(40, 41)^. However, importantly, when these false negative cases were explored in depth, the risk of missing a VTE was deemed low and any adverse event was not thought related to the VTE.

The AAT’s higher specificity (71.7% vs. 65.2%) suggests it is more effective in reducing false positives. Approximately 65.3% of the ST false positive cohort received unnecessary anticoagulation but the clinical review concluded that the haemorrhagic events seen in a subset of these patients were not related to their anticoagulation therapy. However, these patients did go on to receive imaging and had a longer length of stay than the true negatives.

The data demonstrates the diminishing effectiveness of both D-dimer thresholds with advancing age, but more so with the ST. The analysis also showed that the AAT maintains greater accuracy and specificity across various patient subgroups, including those with comorbidities. However, due to small subgroup sample sizes, these findings should be interpreted cautiously.

A significant finding of the study is the potential reduction in unnecessary imaging procedures by adopting the AAT. The analysis suggests a substantial decrease in the number of scans required, resulting in considerable cost savings, a critical consideration for healthcare systems like the NHS.

The results advocate for adopting AAT in clinical settings, especially in populations where age and comorbidities may impact the accuracy of ST, including in a diverse urban setting. Clinicians should weigh the benefits of reduced false positives and resource savings against the slight decrease in sensitivity.

### Strengths and limitations

The major strength of the study was the large sample size, which allowed for analyses of D-dimer within patient subgroups. However, the retrospective study design also led to several limitations, which need to be considered when interpreting the results. Primarily, it was assumed that the ordering of a D-dimer test indicated that a clinician suspected that a patient had VTE. However, the fact that the rate of VTE diagnosis in the present study was considerably lower than similar studies in the literature would suggest that there was a degree of over-testing, where D-dimer tests were ordered in patients at low risk of VTE^(12, 42, 43)^. Consequently, the results of the analysis are only generalisable to situations which apply similar criteria for ordering D-dimer testing and may not be applicable to situations where D-dimer tests are ordered more sparingly. Secondly, the retrospective data collection resulted in some missing data. This was a particular issue for the Wells’ Scores, where the Wells-PE was recorded for only 14.4% of cases, with the Wells-DVT available for <0.1%. Further review found that, whilst these scores were routinely calculated for patients, they were generally recorded in the handwritten patient notes rather than the EHR, and so could not be readily extracted for analysis. As such, it was not possible to perform any meaningful analysis of the Wells-DVT score, and analyses of the Wells-PE must be interpreted with caution, due to the risk of selection bias. Similarly, the presenting complaint was not recorded for patients who attended via either a GP referral or the community DVT pathway; hence, these patients were either excluded or grouped into a “not recorded” category for analysis. This will have introduced selection bias into analyses of the presenting complaint, which is demonstrated by the fact that cases where this was not recorded had the highest rate of VTE diagnosis. Finally, the D-dimer assay used during the study period had a lower limit of 150μg/L, with over 40% of patients being in this range. All of these patients were assigned the same value for analysis, which may have impacted the calculated predictive accuracy of the D-dimer test when analysed as a continuous variable (e.g. when calculating AUROCs).

## Conclusion

In conclusion, this study provides evidence that age-adjusted D-dimer thresholds offer a more accurate and resource-efficient approach for diagnosing VTE, particularly in older patients and those with comorbidities. Adopting this approach could lead to better patient outcomes and significant cost savings, although careful consideration of its limitations and further validation is necessary. This discussion aims to stimulate further research and debate on the optimal use of D-dimer testing in clinical practice, considering the complex interplay of accuracy, patient demographics, and healthcare resource management.

## Data Sharing Agreement

To facilitate knowledge in this area, the anonymised participant data and a data dictionary defining each field will be available to others through application to PIONEER via the corresponding author.

## Data Availability

https://web.www.healthdatagateway.org/dataset/f21ef3fd-8193-447f-bcda-191243ca4b12

## Author contribution

S. Gallier, C. Atkin, B. Holloway, W. Lester and E. Sapey designed the study, collated data, performed some analysis, and wrote the manuscript. F. Evison curated data and supported statistical analysis. R. Khosla undertook initial literature review and assisted in writing the introduction. S. Gallier, L. Rickard, T. Ranasinghe, B. Holloway, V. Reddy-Kolanu, W. Lester and E. Sapey undertook the service evaluation and reviewed the clinical notes. J. Hodson performed statistical analyses and support. S. Gallier performed statistical analysis and wrote the first draft of the manuscript. All authors amended the manuscript and approved the final version.

## Acknowledgements

This work was supported by PIONEER, the Health Data Research Hub in acute care, NIHR Midlands Patient Safety Research Collaboration (PSRC) and the NIHR Applied Research Collaboration (ARC) West Midlands. This work uses data provided by patients and collected by the NHS as part of their care and support. We would like to acknowledge the contribution of all staff, key workers, patients and the community who have supported our hospitals and the wider NHS at this time.

