## Supplementary Table 1 for "The safety and efficacy of using age-adjusted D-dimers in hospitalised patients in a diverse urban centre: a real-world data study"

### Supplementary Analysis

***Supplementary Table 1 - Classification accuracy of D-dimer thresholds by age***

| **Age**  **(Years)** | **D-Dimer Threshold** | **VTE** | | **Sensitivity** | **Specificity** | **NPV** | **PPV** | **Accuracy** |
| --- | --- | --- | --- | --- | --- | --- | --- | --- |
|  |  | ***No*** | ***Yes*** |  |  |  |  |  |
| **<50**  *(N=12240)* |  |  |  | *p=N/A* | *p=N/A* | *p=N/A* | *p=N/A* | *p=N/A* |
|  | ST |  |  |  |  |  |  |  |
|  | *<250μg/L* | 9516 | 39 | 87.5% | 79.8% | 99.6% | 10.1% | 80.0% |
|  | *≥250μg/L* | 2413 | 272 | (272/311) | (9516/11929) | (9516/9555) | (272/2685) | (9788/12240) |
|  | AAT |  |  |  |  |  |  |  |
|  | *<AAT* | 9516 | 39 | 87.5% | 79.8% | 99.6% | 10.1% | 80.0% |
|  | *≥AAT* | 2413 | 272 | (272/311) | (9516/11929) | (9516/9555) | (272/2685) | (9788/12240) |
| **50-59**  *(N=4748)* |  |  |  | *p=0.695* | ***p=0.003*** | *p=0.810* | *p=0.497* | ***p=0.005*** |
|  | ST |  |  |  |  |  |  |  |
|  | *<250μg/L* | 3203 | 33 | 84.5% | 70.6% | 99.0% | 11.9% | 71.3% |
|  | *≥250μg/L* | 1332 | 180 | (180/213) | (3203/4535) | (3203/3236) | (180/1512) | (3383/4748) |
|  | AAT |  |  |  |  |  |  |  |
|  | *<AAT* | 3330 | 37 | 82.6% | 73.4% | 98.9% | 12.7% | 73.8% |
|  | *≥AAT* | 1205 | 176 | (176/213) | (3330/4535) | (3330/3367) | (176/1381) | (3506/4748) |
| **60-69**  *(N=3973)* |  |  |  | *p=0.169* | ***p<0.001*** | *p=0.342* | *p=0.084* | ***p<0.001*** |
|  | ST |  |  |  |  |  |  |  |
|  | *<250μg/L* | 2206 | 10 | 94.9% | 58.4% | 99.5% | 10.7% | 60.3% |
|  | *≥250μg/L* | 1569 | 188 | (188/198) | (2206/3775) | (2206/2216) | (188/1757) | (2394/3973) |
|  | AAT |  |  |  |  |  |  |  |
|  | *<AAT* | 2537 | 18 | 90.9% | 67.2% | 99.3% | 12.7% | 68.4% |
|  | *≥AAT* | 1238 | 180 | (180/198) | (2537/3775) | (2537/2555) | (180/1418) | (2717/3973) |
| **70-79**  *(N=3545)* |  |  |  | ***p=0.017*** | ***p<0.001*** | *p=0.202* | ***p=0.011*** | ***p<0.001*** |
|  | ST |  |  |  |  |  |  |  |
|  | *<250μg/L* | 1474 | 17 | 93.1% | 44.7% | 98.9% | 11.1% | 48.0% |
|  | *≥250μg/L* | 1826 | 228 | (228/245) | (1474/3300) | (1474/1491) | (228/2054) | (1702/3545) |
|  | AAT |  |  |  |  |  |  |  |
|  | *<AAT* | 1998 | 34 | 86.1% | 60.5% | 98.3% | 13.9% | 62.3% |
|  | *≥AAT* | 1302 | 211 | (211/245) | (1998/3300) | (1998/2032) | (211/1513) | (2209/3545) |
| **80-89**  *(N=2393)* |  |  |  | ***p=0.001*** | ***p<0.001*** | *p=0.102* | ***p=0.010*** | ***p<0.001*** |
|  | ST |  |  |  |  |  |  |  |
|  | *<250μg/L* | 662 | 5 | 97.0% | 29.7% | 99.3% | 9.4% | 34.4% |
|  | *≥250μg/L* | 1564 | 162 | (162/167) | (662/2226) | (662/667) | (162/1726) | (824/2393) |
|  | AAT |  |  |  |  |  |  |  |
|  | *<AAT* | 1210 | 22 | 86.8% | 54.4% | 98.2% | 12.5% | 56.6% |
|  | *≥AAT* | 1016 | 145 | (145/167) | (1210/2226) | (1210/1232) | (145/1161) | (1355/2393) |
| **90+**  *(N=627)* |  |  |  | *p=0.617* | ***p<0.001*** | *p=1.000* | *p=0.084* | ***p<0.001*** |
|  | ST |  |  |  |  |  |  |  |
|  | *<250μg/L* | 116 | 1 | 97.8% | 20.0% | 99.1% | 8.8% | 25.7% |
|  | *≥250μg/L* | 465 | 45 | (45/46) | (116/581) | (116/117) | (45/510) | (161/627) |
|  | AAT |  |  |  |  |  |  |  |
|  | *<AAT* | 286 | 3 | 93.5% | 49.2% | 99.0% | 12.7% | 52.5% |
|  | *≥AAT* | 295 | 43 | (43/46) | (286/581) | (286/289) | (43/338) | (329/627) |

***Legend****. The age-adjusted threshold used a value of 250μg/L for those aged <50 years, or age (in years)*5μg/L for those older than 50 years. p-Values are from Fisher’s exact tests, comparing the percentages between the standard and age-adjusted thresholds within each subgroup of age; bold p-values are significant at p<0.05. AAT: Age-Adjusted Threshold, N(P)PV: Negative (Positive) Predictive Value, ST: Standard Threshold, VTE: Venous Thromboembolism.*
