## Supplementary Figure 1 for "The safety and efficacy of using age-adjusted D-dimers in hospitalised patients in a diverse urban centre: a real-world data study"

### Supplementary Analysis

**Supplementary Figure 1 – Study flowchart**

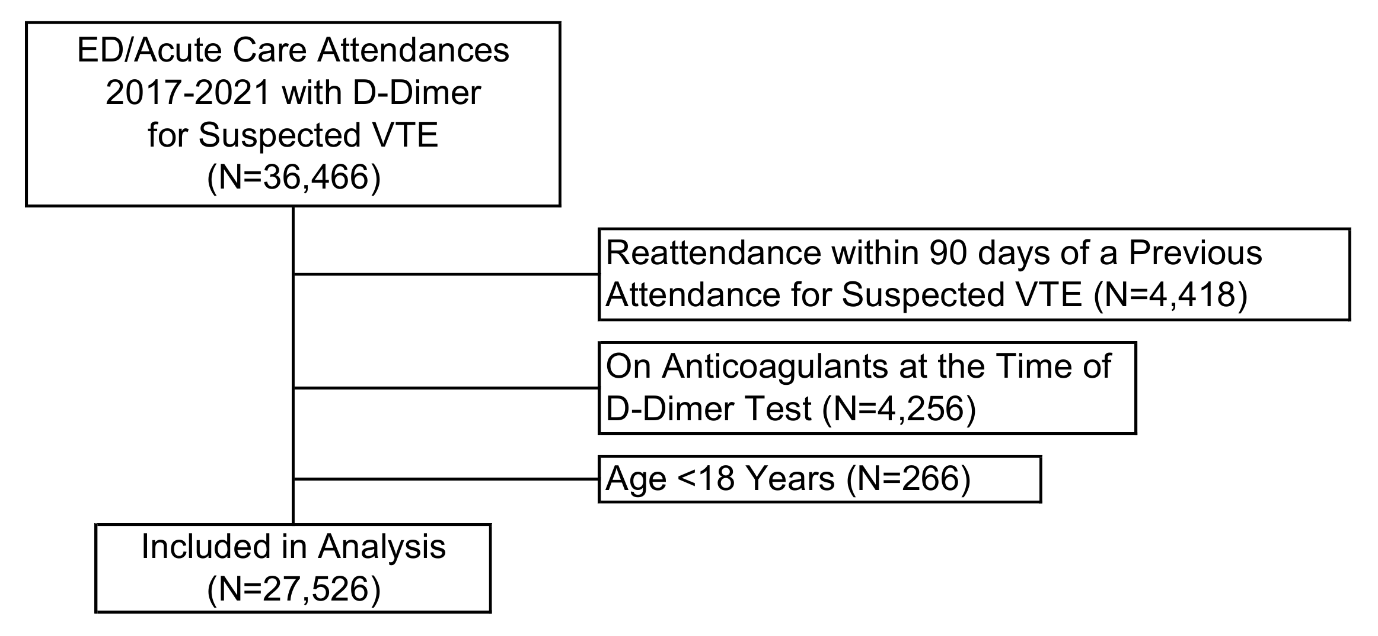

***Legend.*** *ED: Emergency Department, VTE: Venous Thromboembolism*
